# Knowledge, attitudes and willingness to donate gametes among university students in Ghana: a cross-sectional study

**DOI:** 10.64898/2026.09.30.26364451

**Authors:** Michael Y. Amoh, Adu Appiah-Kubi, Anthony K. Dah, Wisdom K. Azanu, Wisdom M. Boateng, Emelda D. Wuaku, Bernard K. Okai, Charles M. Senaya, Rex M. K. Djokoto

## Abstract

Gamete donation may widen access to assisted reproductive technology, but donor gametes remain difficult to obtain in Ghana. This institution-based cross-sectional survey assessed knowledge, attitudes and willingness to donate among 380 undergraduate students at Ho Technical University. The sample was allocated proportionately across 24 strata defined by six faculties and four academic levels; within each stratum, students who were present during scheduled classes were recruited consecutively. Data were collected using a self-administered questionnaire and analysed using descriptive statistics, exact tests and exploratory logistic regression. Although 88.2% of participants had heard of gamete donation and 72.4% supported it, only 42.1% said they would donate if approached. Among the 220 participants who were unwilling, 210 (95.5%) selected the response that donation ‘felt wrong’ to them. Participants who perceived societal attitudes as positive had higher odds of reported willingness than those who perceived them as negative (adjusted odds ratio [aOR] 3.08, 95% confidence interval [CI] 1.34–7.08). Belief that donation could exhaust one’s gametes was associated with lower odds of willingness (aOR 0.27, 95% CI 0.15–0.48). The conditional statement that payment would increase willingness was also associated with direct willingness (aOR 7.16, 95% CI 4.25–12.06), but the two measures overlapped conceptually. The findings identify fertility misconceptions, concerns about donation and differing views on compensation that should be considered when developing donor information, counselling and protection measures.

## Introduction

The World Health Organization defines infertility as failure to achieve pregnancy after 12 months or more of regular unprotected sexual intercourse [1]. Across sub-Saharan Africa, its effects often extend well beyond the clinical diagnosis. Women and couples may experience stigma, marital strain, social exclusion and substantial psychological distress [2–7]. Ghanaian studies describe similar pressures, including strong expectations to have children and the uncertainty faced by couples seeking treatment [4–6].

Assisted reproductive technology (ART), including in vitro fertilisation using donated oocytes or sperm, offers an important treatment option for some people affected by infertility [8]. Access depends not only on the availability of fertility clinics but also on cost, trust, ethical concerns and the supply of donor gametes. In Ghana, ART services have developed over several decades, but they remain concentrated in a small number of centres and are unaffordable for many families [9,10,12]. Donor availability, informed decision-making and protection of donors are therefore relevant to equitable access to ART.

Gamete donation raises questions about lineage, identity, inheritance and family belonging [4,10]. These concerns may carry particular weight in settings where biological parenthood is closely tied to social status and continuity of the family line. Ghanaian qualitative research found that university students could value donation as a way of helping people with infertility while also expressing concerns about religion, secrecy, incest and the social position of donor-conceived children [13]. Support for donation in principle may therefore coexist with reluctance to donate personally.

The findings were interpreted with reference to selected constructs of the Theory of Planned Behaviour, including attitudes, perceived social norms and perceived behavioural control [14]. These constructs were relevant to variables such as personal support for donation, perceived societal attitudes, fertility-related beliefs, risk awareness and views on payment. A validated Theory of Planned Behaviour instrument was not used, and the study was not designed as a formal test of the theory.

Studies from Ghana and Nigeria suggest a gap between awareness of gamete donation and willingness to donate. Among non-medical tertiary students in Ghana, awareness of ART was fairly high, but understanding and acceptability were uneven [15]. Ghanaian qualitative work has likewise shown that students may regard donation as helpful while still expressing religious, sociocultural and family concerns [13]. In Lagos, 96.3% of surveyed undergraduates had heard of gamete donation, but 38.7% had good knowledge and 37.5% were willing to participate [16]. Among female undergraduates in Rivers State, awareness did not consistently translate into willingness, and payment influenced the views of some potential donors [17]. Studies outside Africa have also linked donation decisions to altruism, expectations, awareness of side effects and the practical demands of the procedure [11,18– 20]. Another Nigerian study reported broad support for assisted reproduction but lower willingness to join a donor programme, with ethical concerns common among those who opposed donation [21].

These studies suggest that awareness alone does not explain whether a young adult would donate. Important gaps remain in the Ghanaian evidence. Existing work has focused mainly on general awareness of assisted reproduction or qualitative accounts of egg and sperm donation. Quantitative studies that include male and female students, distinguish general support from personal willingness, and examine social perceptions, fertility misconceptions, risk awareness and views on payment in the same analysis remain limited. Evidence from Nigeria is useful for comparison, but differences in social context, institutional setting and measurement limit direct application to Ghana. This study therefore assessed knowledge, attitudes and willingness to donate gametes among students at Ho Technical University and examined factors associated with willingness to donate if approached.

## Methods

### Study design and study site

An institution-based cross-sectional survey was conducted from July to September 2025 at Ho Technical University in the Volta Region of Ghana. The survey assessed knowledge, attitudes and willingness to donate gametes and explored factors associated with willingness to donate if approached. The university comprises six faculties: Applied Sciences and Technology, Engineering, Art and Design, Built and Natural Environment, Business, and Applied Social Sciences. Male and female undergraduates in Levels 100 to 400 were eligible. Students younger than 18 years could participate under the approved protocol with assent and written parental or guardian consent. Those who declined, withdrew or returned substantially incomplete questionnaires were excluded. Reporting followed the Strengthening the Reporting of Observational Studies in Epidemiology guidance for cross-sectional studies [22].

### Sample size and sampling

The sample size was calculated using the Yamane formula, n = N/[1 + N(e^2^)], where n is the required sample size, N is the study population and e is the margin of error [23]. Based on an undergraduate population of 7,033 and a 5% margin of error, the minimum sample was 378.47 and was rounded to 380. No separate allowance was made for non-response. The sample was allocated proportionately across 24 strata defined by six faculties and four academic levels. Applied Sciences and Technology contributed 130 participants, Engineering 92, Art and Design 59, Built and Natural Environment 40, Business 32 and Applied Social Sciences 27. By academic level, 113 participants were in Level 100, 96 in Level 200, 86 in Level 300 and 85 in Level 400. Within each faculty-by-level stratum, students who were present during scheduled classes were recruited consecutively until the allocated number was reached (Figure 1). This was non-probability consecutive recruitment within predefined strata rather than random selection from the full undergraduate population. The numbers approached, declining participation and returning substantially incomplete questionnaires were not recorded, so a formal response rate and the extent of non-response bias could not be calculated. Proportional allocation improved coverage across faculties and academic levels but did not remove the risk of selection bias.

**Figure 1:**
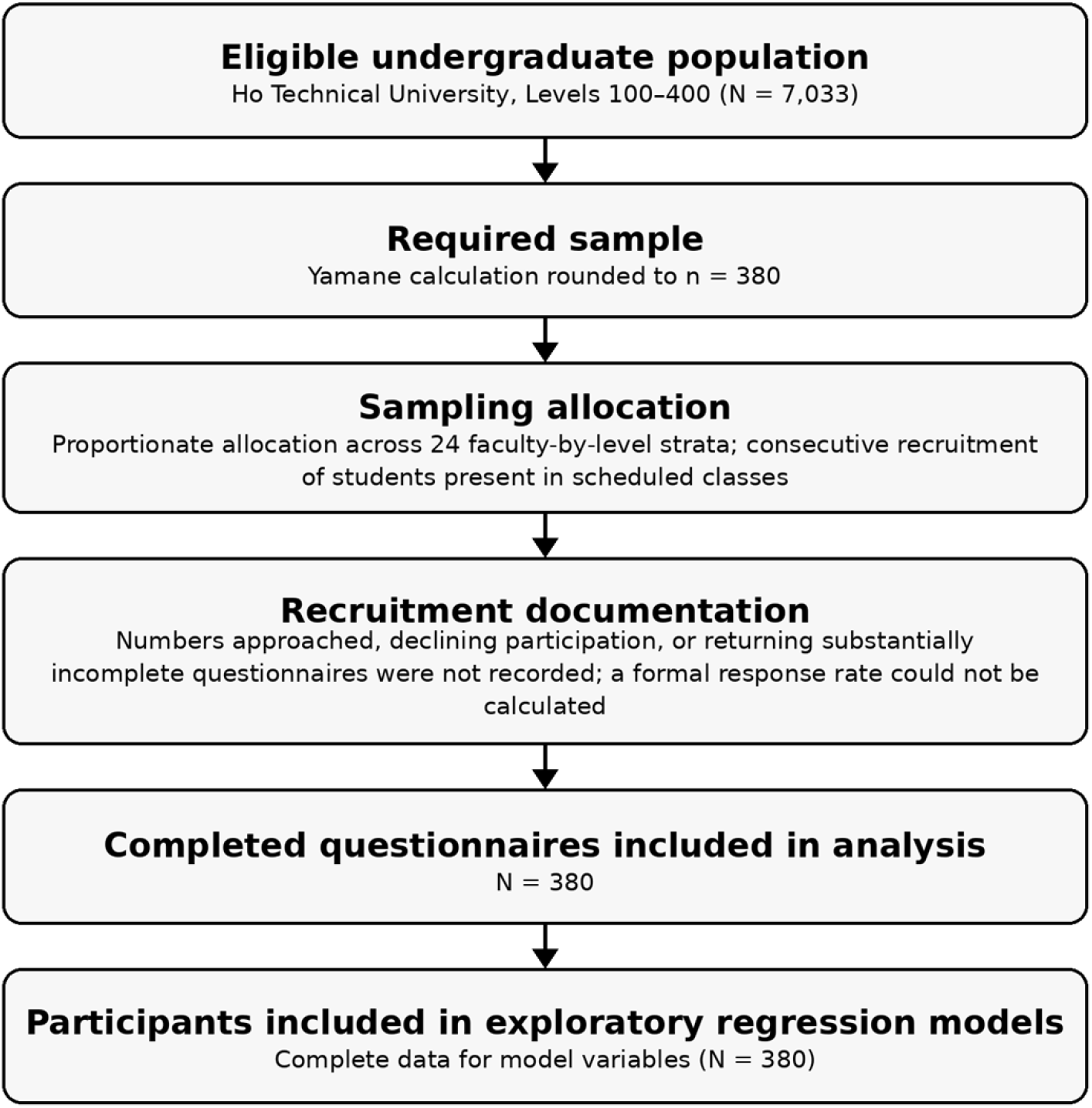
Flow of participants through recruitment and analysis.

### Data collection

A structured self-administered questionnaire was adapted from earlier studies of ART, oocyte donation, sperm donation and reproductive health knowledge, attitudes and practices [13,15,21]. It covered sociodemographic characteristics, awareness and knowledge of ART and gamete donation, attitudes, willingness, motivations and perceived barriers. The questionnaire was pretested among 15 undergraduates at the University of Health and Allied Sciences in Ho who did not take part in the main study. The pretest assessed clarity, relevance and question order, after which minor wording changes were made without altering the intended constructs. It was not a formal psychometric validation of the questionnaire in this population. Participants completed the questionnaire anonymously and were informed that participation would not affect their academic standing. Multiple responses were permitted for reasons for non-support and unwillingness. The response option used to capture moral discomfort was worded as ‘It feels wrong to me’.

### Variables and scoring

The primary direct outcomes were awareness of gamete donation, support for gamete donation and willingness to donate if approached, each coded as yes or no. Exploratory composite indices were also created as secondary measures. Seven knowledge items were scored 1 for a correct response and 0 for an incorrect or ‘do not know’ response, with scores of 4–7 classified as good knowledge. The nine-item attitude-readiness index awarded one point for supportive responses and reverse-scored two fertility misconceptions; scores of 5–9 were classified as positive readiness. Five willingness-related items were scored similarly, with scores of 3–5 classified as positive readiness. These midpoint cut-offs were not derived from a validated instrument, so all three indices were treated as exploratory. Supplementary File 1 provides the items, coding rules and score ranges. The donor-anonymity and known-donor items (Q21 and Q22) were analysed separately because neither has a single clearly favourable direction. Q32, which asked whether the participant wanted information about the recipient family, was retained in the exploratory willingness-readiness index because it formed part of the original scoring plan. Because this index had poor internal consistency, it was used only as an exploratory descriptive measure and not for substantive inference. Explanatory variables included sex, age group, marital status, religion, academic level and faculty. Perception-related variables included awareness of ART, participants’ perception of societal attitudes, beliefs about infertility and gamete exhaustion, anonymity preferences, awareness of donation-related risks and conditional willingness to donate if paid.

### Statistical analysis

Data were checked, coded and entered into Microsoft Excel before analysis using IBM SPSS Statistics version 20. Categorical variables are presented as frequencies and percentages, and continuous variables as means and standard deviations. Chi-square tests were used to examine associations between sociodemographic characteristics and dichotomised outcomes. Fisher’s exact test was used for sparse 2 × 2 tables, and the Fisher–Freeman–Halton exact test for larger tables when more than 20% of expected cell counts were below 5. Logistic regression was exploratory. Perception-related variables associated with direct willingness at p<0.05 in bivariate analysis were entered into the adjusted model, while sex was retained a priori. Adjusted odds ratios (aORs) and 95% confidence intervals were reported, with statistical significance set at p<0.05. No adjustment was made for multiple comparisons, so isolated bivariate findings were interpreted cautiously. Model fit was assessed using the Hosmer–Lemeshow test. A sensitivity analysis repeated the model without the payment variable while retaining sex, participants’ perception of societal attitudes, belief that donation causes infertility and belief that donation exhausts one’s gametes. All 380 participants had complete data for the variables included in the regression models. No prespecified subgroup analyses or interaction tests were performed. Internal consistency was assessed using the Kuder–Richardson Formula 20 (KR-20), with coefficients of 0.65 for knowledge, 0.80 for attitude-readiness and 0.31 for willingness-readiness. The willingness-readiness index was therefore reported only as an exploratory descriptive measure. Direct willingness remained the primary willingness outcome and was used in the regression analysis.

### Ethical considerations

Ethical approval was obtained from the Research Ethics Committee of the University of Health and Allied Sciences, Ghana (UHAS-SOM REC A/134/2024-2025), and administrative permission was obtained from Ho Technical University. All adult participants provided written informed consent. The two participants aged 17 years provided assent together with written parental or guardian consent, as required by the approved protocol. Participation was voluntary, and participants could withdraw at any time without penalty. Questionnaires were anonymised using study codes, and access to stored data was restricted to the research team.

## Results

### Participant characteristics

All 380 completed questionnaires were included in the analysis, meeting the calculated minimum sample size (Table 1). The mean age was 21.93 years (standard deviation 2.41). Most participants were aged 21–25 years (58.9%), just over half were male (52.4%), and the great majority were single (95.8%) and Christian (95.0%).

**Table 1:**
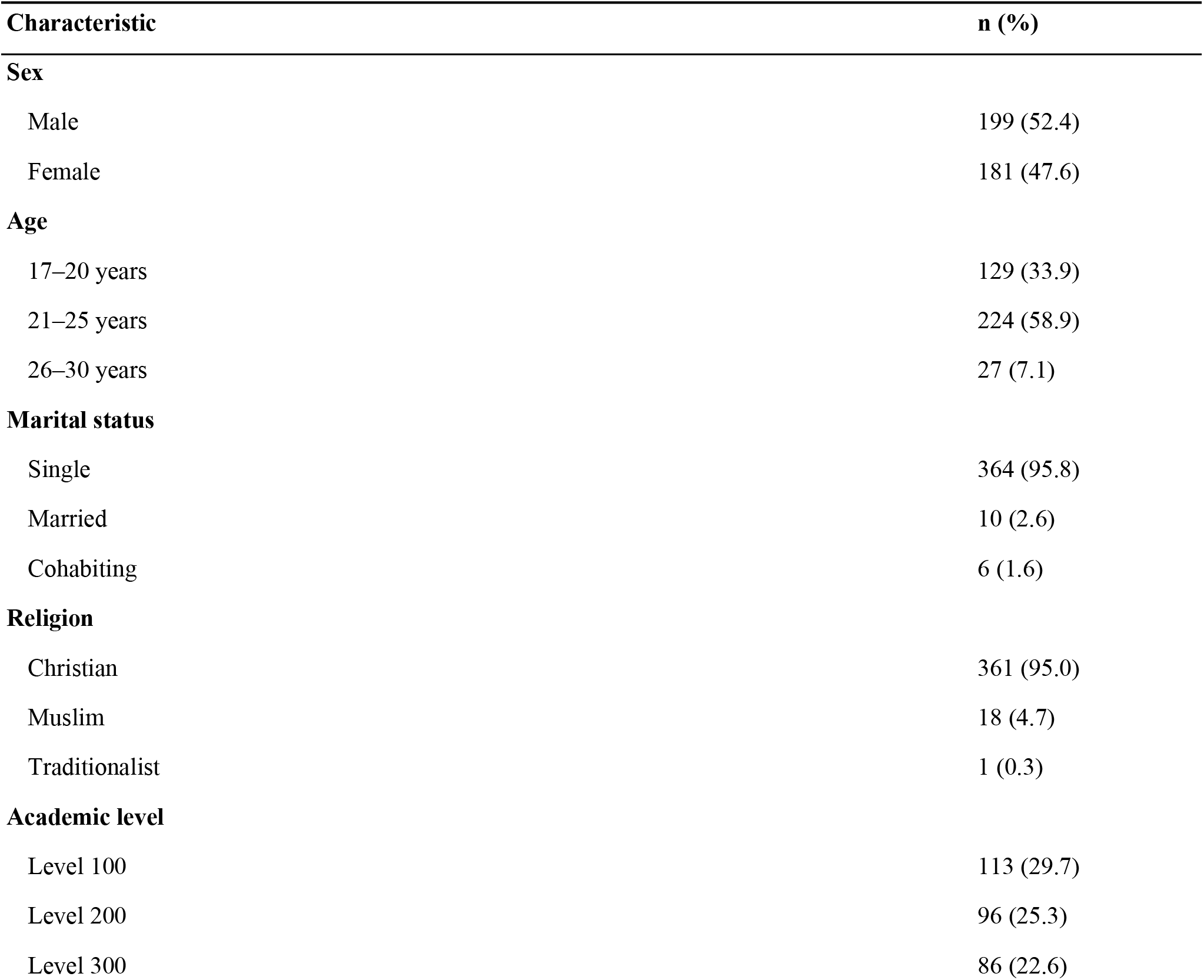
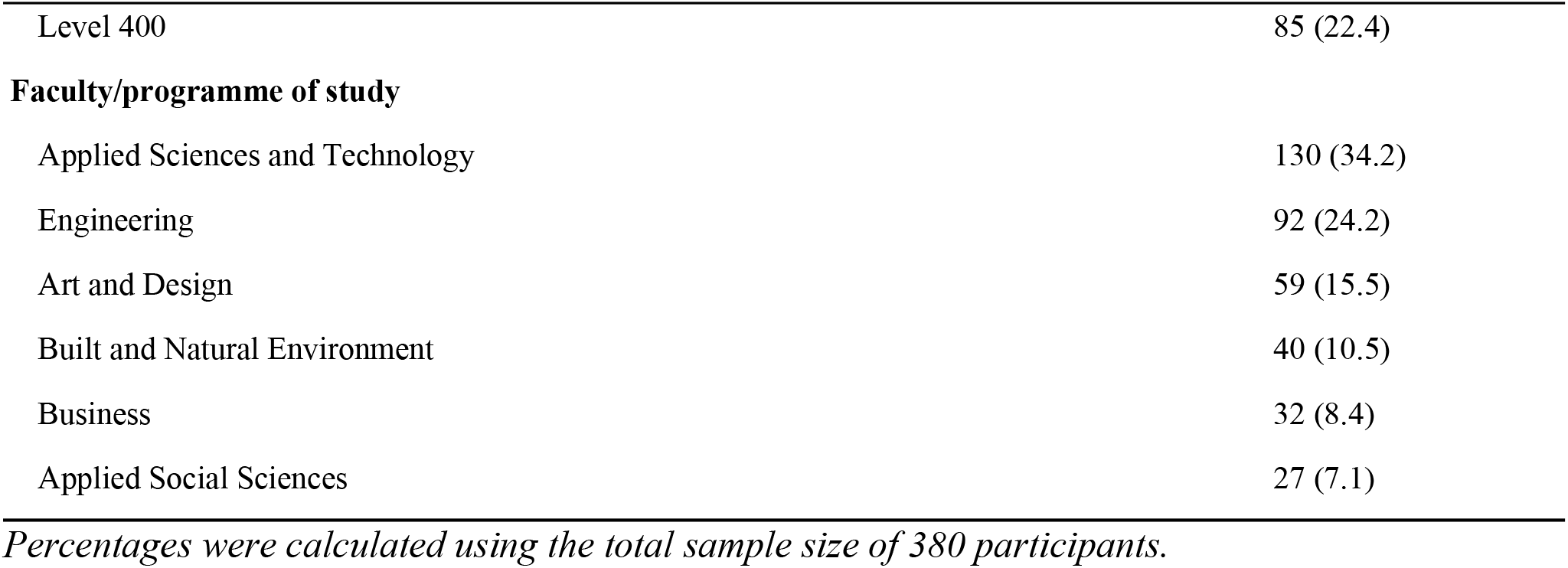
Sociodemographic characteristics of participants (N=380)

### Knowledge and awareness

Awareness of gamete donation and assisted reproductive technology (ART) was high: 88.2% of participants had heard of gamete donation (95% CI 84.5–91.0%) and 87.9% had heard of ART. Overall, 68.4% recognised gamete donation as a form of ART, while 75.5% knew that ART was practised in Ghana. Knowledge of available services was more limited. Only 20.3% knew of a hospital or clinic in Ghana where gamete donation was practised, and 5.5% knew of such a facility in Ho. Based on the exploratory knowledge index, 57.1% were classified as having good knowledge. In addition, 39 participants (10.3%) personally knew someone who had donated gametes (Table 2).

**Table 2:** Awareness and knowledge of assisted reproductive technology and gamete donation.

| Variable | n (%) |
| --- | --- |
| **Heard of gamete donation** |  |
| Yes | 335 (88.2) |
| No | 45 (11.8) |
| **Heard of assisted reproductive technology** |  |
| Yes | 334 (87.9) |
| No | 46 (12.1) |
| **Aware that gamete donation is a form of assisted reproductive technology** |  |
| Yes | 260 (68.4) |
| No | 120 (31.6) |
| **Aware that assisted reproductive technology is practised in Ghana** |  |
| Yes | 287 (75.5) |
| No | 93 (24.5) |
| **Knows of hospitals or clinics in Ghana where gamete donation is practised** |  |
| Yes | 77 (20.3) |
| No | 303 (79.7) |
| **Knows of hospitals or clinics in Ho where gamete donation is practised** |  |
| Yes | 21 (5.5) |
| No | 359 (94.5) |
| **Knows someone who has donated gametes** |  |
| Yes | 39 (10.3) |
| No | 341 (89.7) |
| **Exploratory knowledge index** |  |
| Good | 217 (57.1) |
| Poor | 163 (42.9) |

ART, assisted reproductive technology. Percentages were calculated using the total sample size of 380 participants.

### Attitudes toward gamete donation

General support for gamete donation was high, although responses to individual attitude items varied. Overall, 72.4% supported gamete donation (95% CI 67.7–76.6%), 70.3% believed that more people should be encouraged to donate, and 68.2% would support a friend or family member who wished to donate. However, 35.3% believed that donation could cause infertility, and the same proportion believed that their gametes could be exhausted through donation. Opinions on anonymity were mixed: 67.6% supported donor-recipient anonymity, while 43.4% believed that donors should be personally known to recipients. The exploratory attitude-readiness index classified 71.3% as having positive readiness, whereas only 10.8% perceived societal attitudes towards gamete donation in Ghana as positive. Among the 105 participants who did not support gamete donation, 62 (59.0%) selected the response that donation ‘felt wrong’ to them, 34.3% cited religious reasons and 15.2% cited cultural reasons. Participants could report more than one reason (Table 3).

**Table 3:**
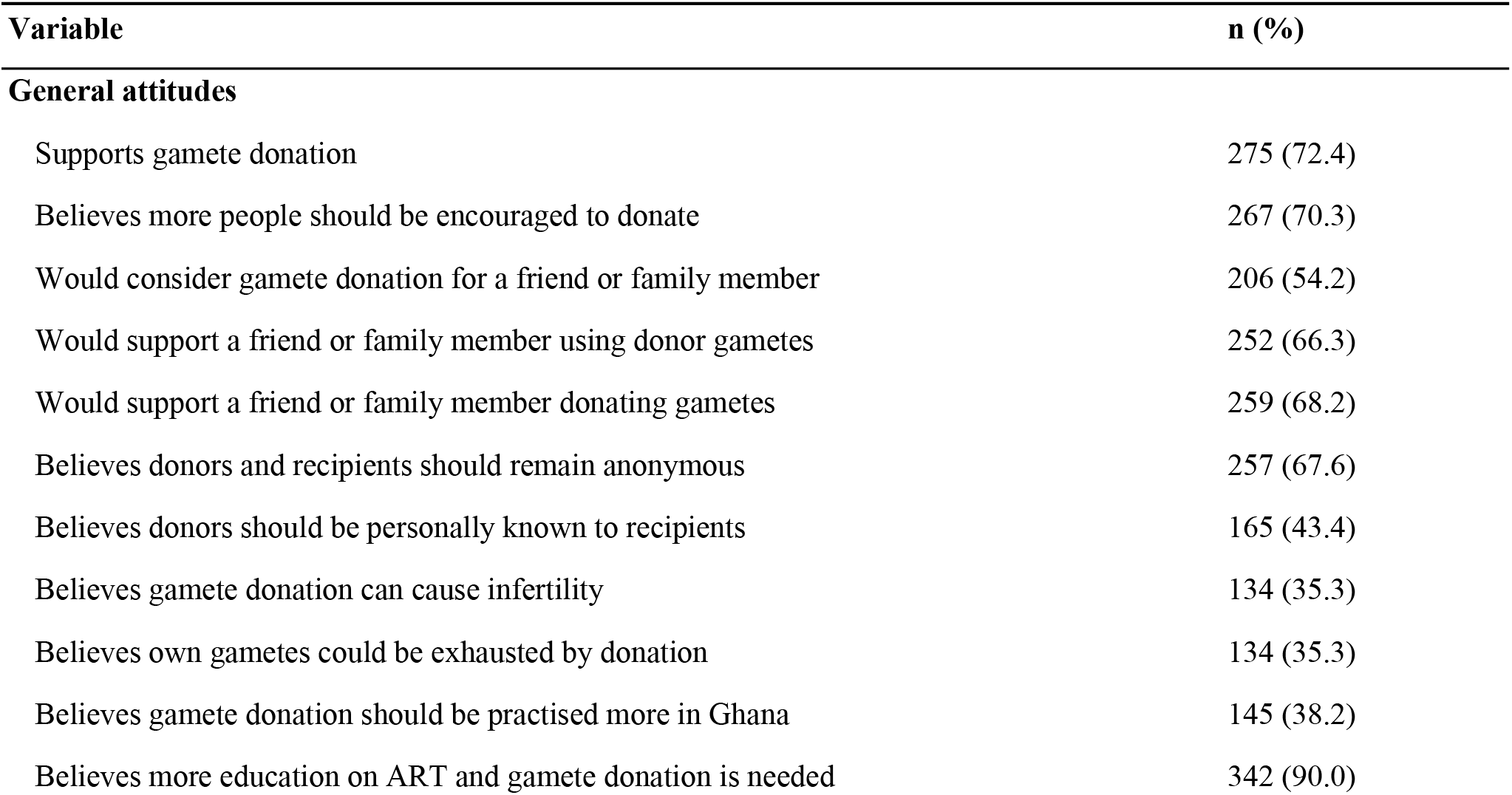
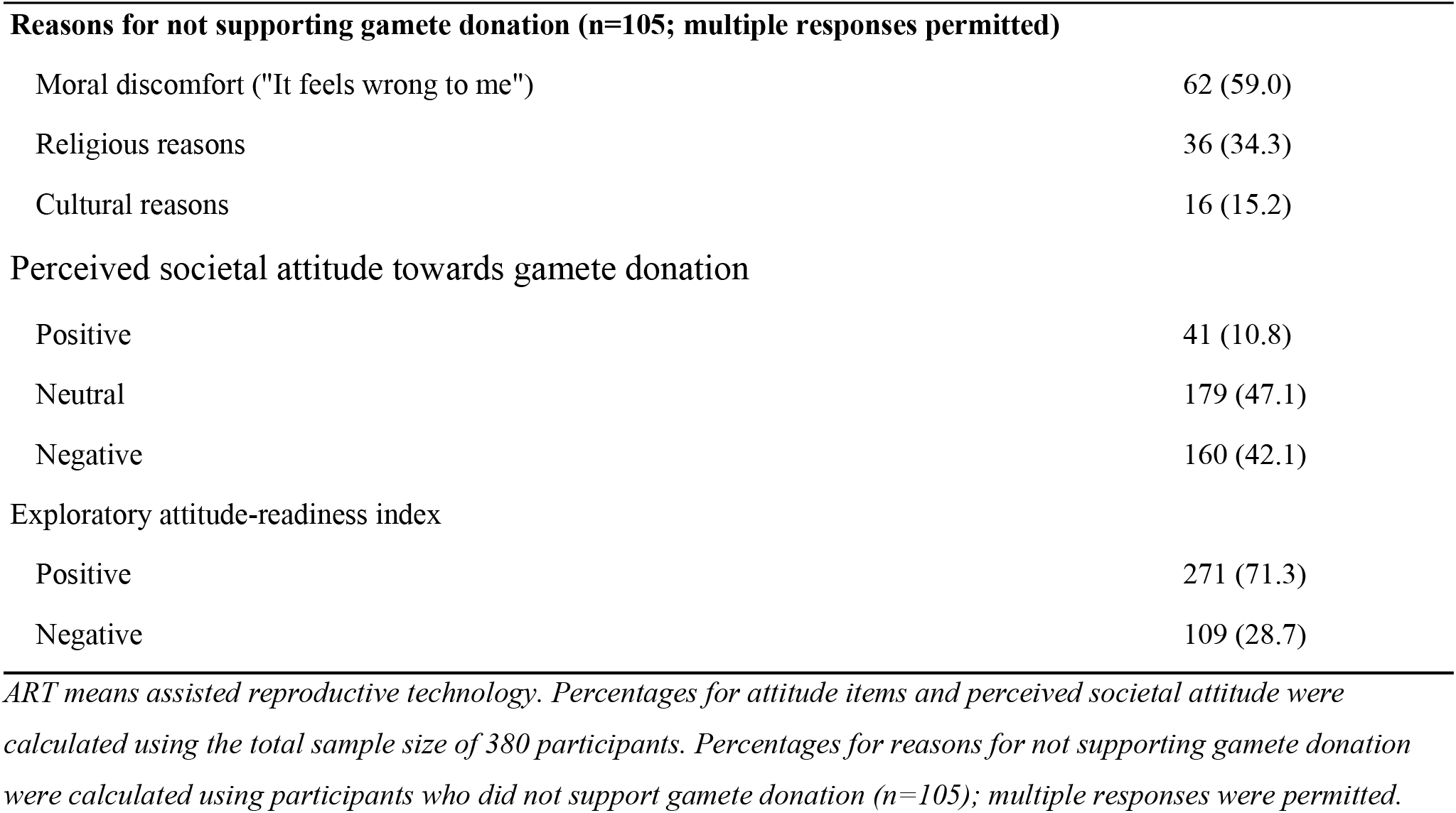
Attitudes towards gamete donation.

| Variable | n (%) |
| --- | --- |
| **General attitudes** |  |
| Supports gamete donation | 275 (72.4) |
| Believes more people should be encouraged to donate | 267 (70.3) |
| Would consider gamete donation for a friend or family member | 206 (54.2) |
| Would support a friend or family member using donor gametes | 252 (66.3) |
| Would support a friend or family member donating gametes | 259 (68.2) |
| Believes donors and recipients should remain anonymous | 257 (67.6) |
| Believes donors should be personally known to recipients | 165 (43.4) |
| Believes gamete donation can cause infertility | 134 (35.3) |
| Believes own gametes could be exhausted by donation | 134 (35.3) |
| Believes gamete donation should be practised more in Ghana | 145 (38.2) |
| Believes more education on ART and gamete donation is needed | 342 (90.0) |
| **Reasons for not supporting gamete donation (n=105; multiple responses permitted)** |  |
| Moral discomfort ("It feels wrong to me") | 62 (59.0) |
| Religious reasons | 36 (34.3) |
| Cultural reasons | 16 (15.2) |
| **Perceived societal attitude towards gamete donation** |  |
| Positive | 41 (10.8) |
| Neutral | 179 (47.1) |
| Negative | 160 (42.1) |
| **Exploratory attitude-readiness index** |  |
| Positive | 271 (71.3) |
| Negative | 109 (28.7) |

*ART means assisted reproductive technology. Percentages for attitude items and perceived societal attitude were calculated using the total sample size of 380 participants. Percentages for reasons for not supporting gamete donation were calculated using participants who did not support gamete donation (n=105); multiple responses were permitted.*

### Willingness to donate

Of the 380 participants, 160 (42.1%) (95% CI 37.2–47.1%) reported that they would donate gametes if approached. The exploratory willingness-readiness index classified 29.7% as having positive readiness; because of its poor internal consistency, it is reported only as a descriptive secondary measure in Supporting Information (S2 Table) rather than in Table 4. Among participants willing to donate, 46.2% identified financial compensation as a motivation, 43.8% reported altruistic motives and 36.2% wished to help a family member or friend. Among the 220 participants who were unwilling, 210 (95.5%) selected the response that donation ‘felt wrong’ to them, 27.7% reported health concerns, 26.8% feared that donation might affect their fertility and 18.2% reported religious reasons. More than half of all participants (54.2%) stated that payment would increase their willingness to donate (Table 4).

**Table 4:** Willingness, motivations and barriers related to gamete donation.

| Variable | Response/category | n (%) |
| --- | --- | --- |
| **Previous donation** |  |  |
| Previous gamete donation | Yes | 15 (3.9) |
| **Direct willingness** |  |  |
| Would donate gametes if approached | Yes | 160 (42.1) |
|  | No | 220 (57.9) |
| **Motivations among participants willing to donate (n=160; multiple responses permitted)** |  |  |
| Reason for willingness | Financial compensation | 74 (46.2) |
|  | Altruism | 70 (43.8) |
|  | Help a family member or friend | 58 (36.2) |
| Barriers among participants unwilling to donate (n=220; multiple responses permitted) |  |  |
| Reason for unwillingness | Moral concerns | 210 (95.5) |
|  | Health concerns | 61 (27.7) |
|  | Fear of affecting fertility | 59 (26.8) |
|  | Religious reasons | 40 (18.2) |
| **Preferences and risk awareness** |  |  |
| Would want to know about the family receiving the donation | Yes | 199 (52.4) |
| More willing to donate if paid | Yes | 206 (54.2) |
| Aware of risks or problems from donating eggs or sperm | Yes | 117 (30.8) |

*Percentages for previous donation, direct willingness, preferences and risk awareness were calculated using the total sample size (N=380). Percentages for motivations were calculated among participants willing to donate (n=160). Percentages for barriers were calculated among participants unwilling to donate (n=220). Multiple responses were permitted for motivations and barriers.*

### Associations with sociodemographic characteristics

No statistically significant associations were found between sex, age group, marital status, academic level or faculty and the exploratory knowledge, attitude-readiness or willingness-readiness indices (all p>0.05). Religion was associated with the exploratory attitude-readiness index using the Fisher– Freeman–Halton exact test (exact p=0.028). This finding is difficult to interpret because 361 participants were Christian, 18 were Muslim and only one was a Traditionalist, resulting in sparse cells, and several bivariate comparisons were conducted. No substantive conclusion about religious differences was drawn. Sex was not associated with direct willingness after adjustment. The remaining sociodemographic variables were not included in the multivariable model.

### Factors associated with willingness to donate

In the bivariate analyses, direct willingness to donate was associated with participants’ perception of societal attitudes, belief that donation could cause infertility, belief that donation could exhaust one’s gametes and the view that payment would increase willingness (all p<0.001). In the adjusted model, participants who perceived societal attitudes as positive had higher odds of reported willingness than those who perceived them as negative (aOR 3.08, 95% CI 1.34–7.08). Belief that donation could exhaust one’s gametes was associated with lower odds of willingness (aOR 0.27, 95% CI 0.15–0.48). Participants who said payment would increase their willingness also had higher odds of direct willingness (aOR 7.16, 95% CI 4.25–12.06). This payment item measured conditional willingness and overlapped conceptually with the primary outcome, so its estimate should not be interpreted as an independent causal or explanatory effect. Belief that donation causes infertility was not statistically significant after adjustment (Table 5). All 380 participants were included in the model, and the Hosmer– Lemeshow test indicated adequate fit (χ2=5.10, df=8, p=0.747). Variance inflation factors for all predictors in the adjusted model ranged from 1.02 to 1.21, indicating no meaningful multicollinearity, and the model showed good discrimination (c-statistic/AUC=0.81). In the sensitivity analysis excluding the payment variable, the associations with participants’ perception of societal attitudes and belief in gamete exhaustion remained in the same direction. Belief that donation causes infertility was statistically significant in this sensitivity model (aOR 0.59, 95% CI 0.36–0.99, p=0.046), although the estimate was sensitive to model specification (Supplementary Table S1).

**Table 5:**
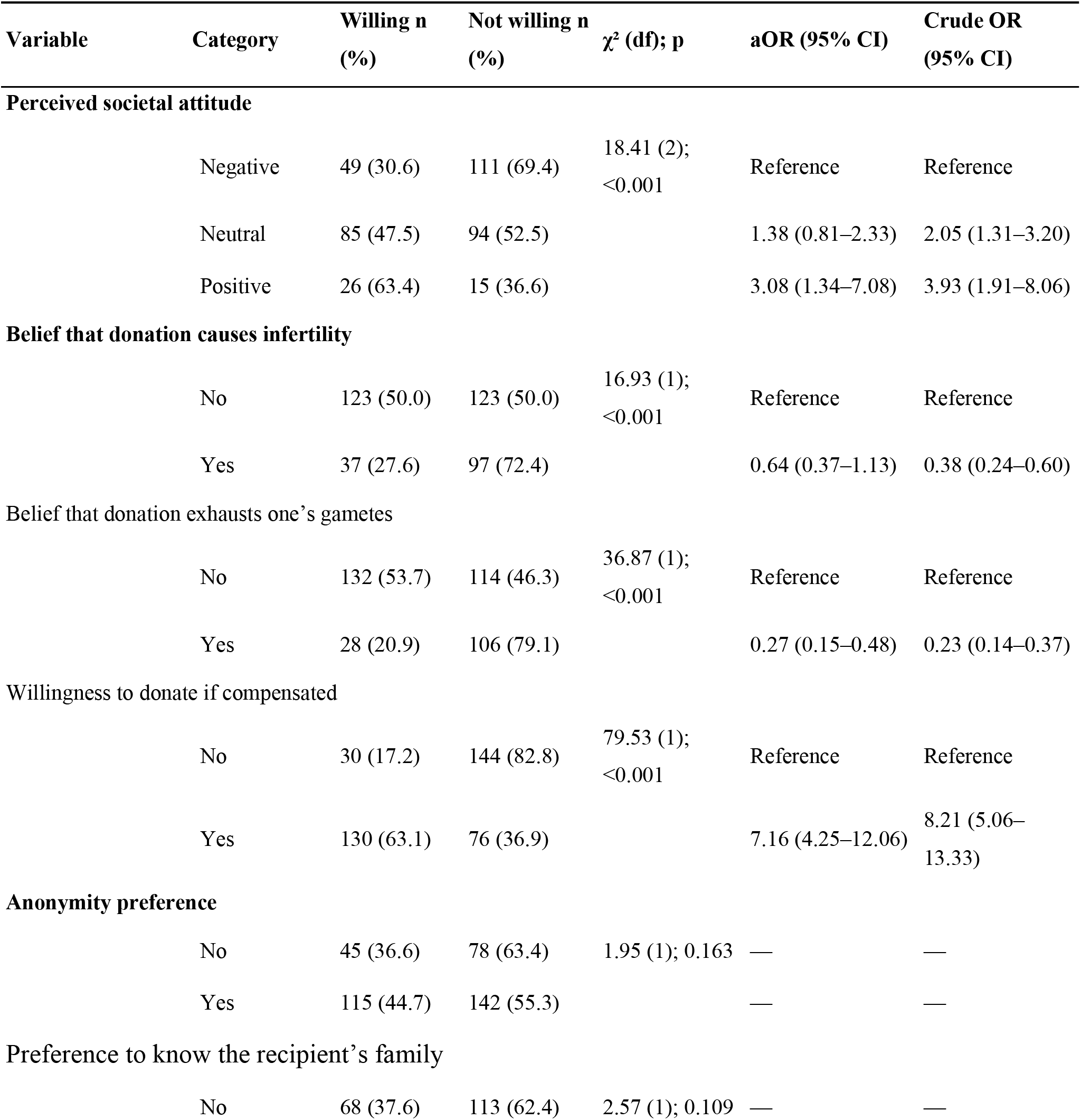
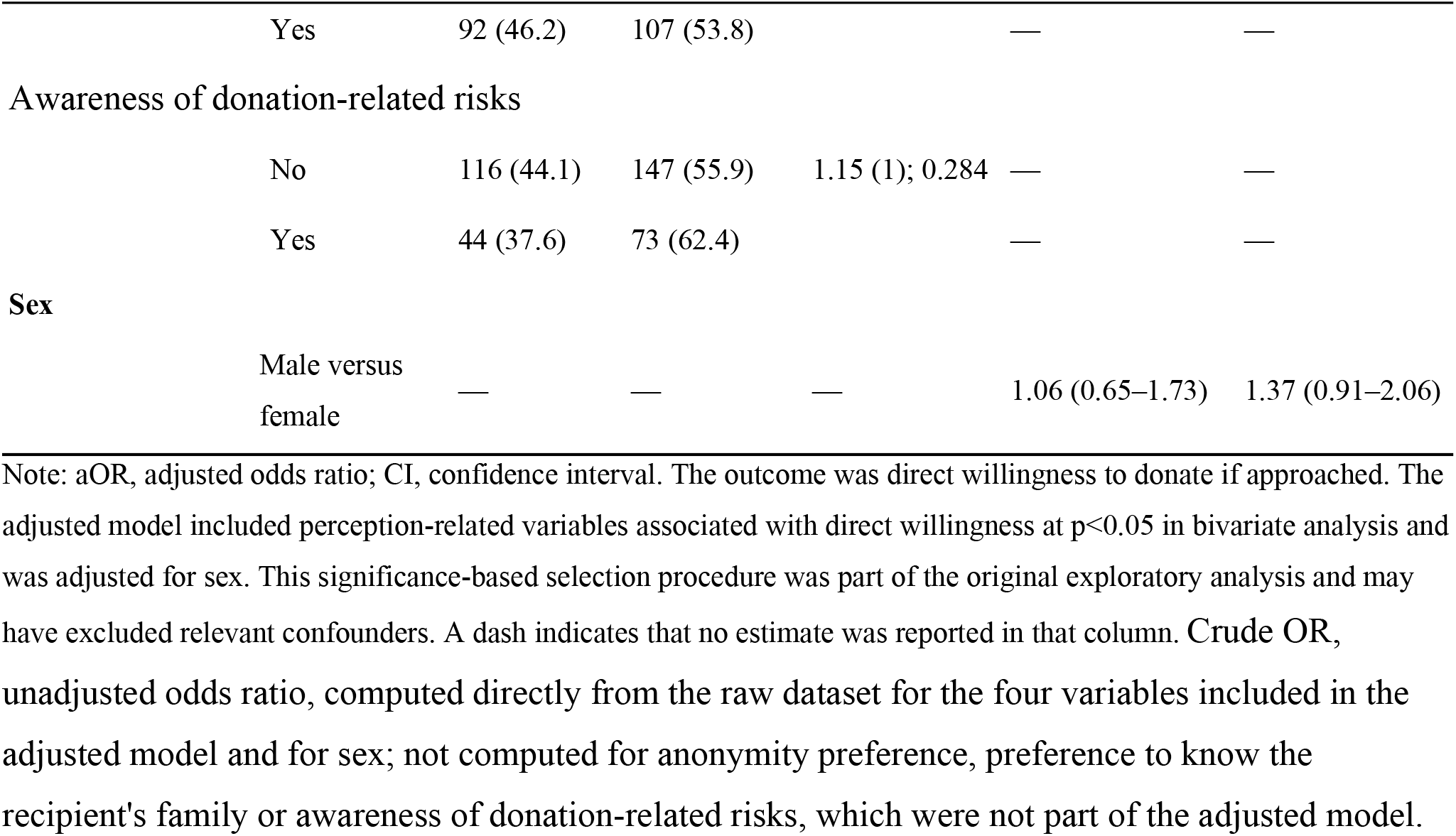
Cross-tabulations and multivariable logistic regression analysis of factors associated with direct willingness to donate gametes.

## Discussion

The main finding was a clear difference between general support for gamete donation and personal willingness to donate. Most participants had heard of gamete donation and supported it, but fewer than half said they would donate if approached. Similar differences have been reported among tertiary students in Ghana and Nigeria, where awareness or approval of ART and gamete donation has not always translated into willingness to participate personally [13,15–17,21]. The present findings suggest that awareness alone does not capture the concerns that shape stated willingness. Awareness was high, but practical knowledge was more limited. Few participants knew where gamete donation was practised, and only 30.8% were aware of possible risks. Hiadzi and Akrong similarly found that Ghanaian university students often knew that donation existed without understanding the procedure in detail [13]. Information provided to potential donors should therefore go beyond defining ART and explain donor screening, the steps involved in sperm and oocyte donation, possible risks, confidentiality, donor rights and where reliable services can be found. Support for donation coexisted with uncertainty about its reproductive effects. More than one-third of participants believed that donation could cause infertility or exhaust their gametes. Similar misconceptions have been reported among Nigerian tertiary students [16,17,21]. These beliefs may affect decisions about donation, particularly among young adults concerned about future fertility.

Accurate information on the procedures and reproductive consequences of sperm and oocyte donation is therefore relevant when potential donors are counselled. Views on anonymity were also divided. Some participants supported anonymous donation, whereas others thought that donors should be known to recipients or wanted information about the recipient family. Ghanaian and international studies have described similar tensions between privacy, kinship and interest in what happens after donation [13,24–27]. A favourable attitude score should therefore not be taken as evidence that a participant is ready to donate.

The response that donation ‘felt wrong’ was the most frequently selected reason among participants who were unwilling to donate, although this item was not included as an independent variable in the regression model. Participants could choose more than one reason, and the wording was broad, so it should not be interpreted as a precise measure of moral or religious objection. In Ghana and other African settings, reproduction may be closely tied to lineage, kinship, inheritance and religious identity [4,6,10,13]. Ghanaian qualitative research has described concerns about incest, secrecy and the use of gametes outside socially recognised family relationships [13]. Nigerian studies have also reported ethical and religious objections alongside otherwise favourable views of ART [16,21]. Information alone may not address all of these concerns; counselling and public discussion may also need to recognise the social meanings attached to donation.

Financial compensation was the most commonly selected motivation among participants who were willing to donate. The conditional statement that payment would increase willingness was also strongly associated with direct willingness after adjustment. A recent Nigerian study among female undergraduates identified financial compensation as a common incentive for potential oocyte donors [17]. The association in the present study requires caution because the payment item overlapped conceptually with the primary outcome. The study did not measure coercion, exploitation or an appropriate level of reimbursement. The finding therefore raises questions for donor information and protection rather than establishing an effect of payment. Any policy discussion should distinguish reimbursement of reasonable expenses from financial inducement and should protect voluntary decision-making [28,29].

### Strengths and limitations

This study included male and female participants from all six faculties and all four academic levels and reached the calculated recruitment target. It adds mixed-sex evidence on a reproductive health topic that has received limited quantitative study in Ghana. The findings nevertheless need careful interpretation. The cross-sectional design cannot establish causation, and stated willingness may not translate into actual donation. Responses may also have been affected by misunderstanding, recall or social desirability. Recruitment was limited to students who were present in class, so students who were absent or unwilling to participate may have held different views. The numbers approached, declining participation and returning incomplete questionnaires were not documented; consequently, neither the response rate nor the extent of non-response bias could be assessed. The findings describe the surveyed participants and should not be treated as population-level estimates for all undergraduates at Ho Technical University or generalised without caution to other Ghanaian institutions or young adults outside tertiary education. The Yamane formula provided a practical recruitment target but was not based on an anticipated effect size or the requirements of the regression model. The questionnaire was adapted from earlier work and pretested for clarity, relevance and flow, but it was not formally validated in this population. It also treated gamete donation broadly and did not adequately distinguish sperm donation from oocyte donation, which differ in procedure, invasiveness, risk, time burden and clinical requirements. Responses to generic questions about gamete donation therefore cannot be assumed to reflect equivalent willingness to donate sperm and oocytes. The composite indices combined heterogeneous items and used unvalidated midpoint cut-offs. The willingness-readiness index had poor internal consistency (KR-20=0.31) and was retained only as an exploratory descriptive measure; direct willingness is the more interpretable outcome. Multiple bivariate comparisons were performed without adjustment, and the isolated religion finding may have occurred by chance. Variable selection based on bivariate significance may have excluded relevant confounders, while the payment variable overlapped conceptually with the outcome. The regression findings should therefore be regarded as exploratory.

## Conclusion

Awareness and general support for gamete donation were common among the surveyed students, but fewer than half reported that they would donate if approached. Among those who were unwilling, many selected the response that donation ‘felt wrong’ to them, and misconceptions about possible effects on fertility were also common. Participants’ perception of societal attitudes and belief that donation could exhaust their gametes were associated with reported willingness. The association between conditional willingness if compensated and direct willingness should be interpreted cautiously because the two measures overlapped conceptually. These findings identify issues that may need to be addressed in donor information, counselling and protection, but they should not be interpreted as population-level estimates or evidence of causal relationships.

## Data Availability

The de-identified dataset underlying the findings of this study is available as Supporting Information file S1 Data (Coded data set).

## Data availability

The de-identified minimal dataset underlying these findings is available from the corresponding author upon reasonable request, subject to approval by the Research Ethics Committee of the University of Health and Allied Sciences (UHAS-SOM REC A/134/2024-2025), because the informed consent obtained from participants did not extend to public deposition of the dataset.

## Acknowledgments

The authors thank the management and students of Ho Technical University for their cooperation during data collection, and the undergraduates at the University of Health and Allied Sciences, Ho, who took part in questionnaire pretesting.

## Supporting information

**S1 File. Questionnaire items, coding rules and score ranges**. Items, coding rules and score ranges for the exploratory knowledge, attitude-readiness and willingness-readiness indices.

**S1 Table. Sensitivity analysis excluding the payment variable**. Multivariable logistic regression model for direct willingness to donate gametes, repeated without the payment variable and retaining sex, perceived societal attitude, belief that donation causes infertility and belief that donation exhausts one’s gametes.

**S2 Table. Exploratory willingness-readiness index**. Distribution of the exploratory willingness-readiness index (positive versus negative readiness), moved here from the main text because of poor internal consistency (KR-20=0.31).

